# Genome-wide association study of COVID-19 Breakthrough Infections and genetic overlap with other diseases: A study of the UK Biobank

**DOI:** 10.1101/2024.08.11.24311845

**Authors:** Yaning Feng, Kenneth Chi-Yin Wong, Wai Kai Tsui, Ruoyu Zhang, Yong Xiang, Hon-Cheong So

**Author notes:** **Correspondence to:** Hon-Cheong So, Lo Kwee-Seong Integrated Biomedical Sciences Building, The Chinese University of Hong Kong, Shatin, Hong Kong.

## Abstract

**Background:** The COVID-19 pandemic has led to substantial health and financial burden worldwide, and vaccines provide hope to reduce the burden of this pandemic. However, vaccinated people remain at risk for SARS-CoV-2 infection. Genome-wide association studies (GWAS) may allow for the identification of potential genetic factors involved in the development of COVID-19 breakthrough infections (BI), however very few or no GWAS have been conducted for COVID-19 BI so far.

**Methods:** We conducted a GWAS and detailed bioinformatics analysis on COVID-19 BI in a European population based on the UK-Biobank (UKBB). We conducted a series of analyses at different levels, including SNP-based, gene-based, pathway, and transcriptome-wide association analyses, to investigate genetic factors associated with COVID-19 BI and hospitalized infection. Polygenic risk score (PRS) and Hoeffding’s test were performed to reveal genetic relationships between BI and other medical conditions.

**Results:** Two independent loci (LD-clumped at r^2^=0.01) reached genome-wide significance (p<5e-08), including rs36170929 mapped to *LOC102725191*/*VWDE,* and rs28645263 mapped to *RETREG1*. Pathway enrichment analysis highlighted pathways such as viral myocarditis, Rho-selective guanine exchange factor AKAP13 signaling, and lipid metabolism. PRS analyses showed significant genetic overlap between COVID-19 BI and heart failure, HbA1c and type 1 diabetes. Genetic dependence was also observed between COVID-19 BI and asthma, lung abnormalities, schizophrenia, and type 1 diabetes, based on the Hoeffding’s test.

**Conclusions:** This GWAS study revealed two significant loci that may be associated with COVID-19 BI, and a number of genes and pathways that may be involved in BI. Genetic overlap with other diseases was identified. Further studies are warranted to replicate the findings and elucidate the mechanisms involved.

## Introduction

COVID-19 has resulted in substantial health and financial burden worldwide. According to the data published by World Health Organization (WHO), over 700 million confirmed cases and 7 million deaths have been reported worldwide as of 1 Jan 2024^1^. Vaccines for COVID-19 are widely perceived to be the most promising strategy to minimize severe disease, mortality, and the burden of this pandemic.

COVID-19 vaccination also reduces risks of infection and transmission, especially prior to the emergence of Omicron variants. In an English study of 151,821 contacts of 99,567 index patients in 2021, the rate of transmission from people fully vaccinated with BNT162b2 (Pfizer-BioNTech) was 23% vs 49% for transmission from unvaccinated people (adjusted odds ratio [aOR], 0.35 [95% CI, 0.26-0.48] for transmission of Delta to unvaccinated contacts; aOR, 0.10 [95%CI, 0.08-0.13] for transmission of Delta to fully vaccinated contacts)^2^.

Nevertheless, evidence shows that fully vaccinated people still remain at risk for SARS-CoV-2 infection. For example, a total of 10,262 SARS-CoV-2 vaccine breakthrough infections had been reported from 46 U.S. states and territories from 1 Jan, 2021 to April 30, 2021^3^, in the period shortly after the launch of vaccination. It is intriguing to study why some individuals are susceptible to breakthrough infection (BI) or severe disease despite vaccination.^3^

Importantly, BI is uncommon in the pre-Omicron period since the vaccine provides a high protection against infection and severe disease^3^; as such, those who indeed develop BI may have specific genetic and/or clinical risk factors. For Omicron variants, vaccination in general provides much weaker protection against infection and the protective effects wanes more quickly. For example, a recent study^4^ of Omicron variants showed that 100 days after immunization, vaccine effectiveness for infection was 26% and 35% for three and four doses of the BioNTech BNT162b2 vaccine, and to 6% and 11% for three and four doses of the CoronaVac inactivated vaccine. Other studies also found low to moderate protective effects and quick waning in the Omicron era^5^. We therefore chose to focus on infection (and severe COVID-19) in the pre-Omicron period; otherwise, we may be *finding genetic variants associated with infections/severe disease in general*, *instead of genetic factors specifically linked to immune responses to vaccination and BI*. Overall, we believe that learning about BI may provide important biological and clinical insights into the pathophysiology of COVID-19 and the immunological mechanisms underlying vaccine responses.

Several studies have been conducted on BI of COVID-19. Sun et al.^6^ identified that persons with immune dysfunction had a substantially higher risk for COVID-19 BI. Bergwerk et al.^7^ conducted a study on BI in healthcare workers, and found that the occurrence of COVID-19 BI was correlated with neutralizing antibody titers during the peri-infection period and most BI were mild or asymptomatic, although persistent symptoms did occur. Kim et al.^8^ presented a case series of vaccinated subjects who were later hospitalized from COVID-19, and found 7 out of 10 patients did not show observed serological response to mRNA vaccination.

However, most studies of COVID-19 BI did not study the influence of genetic factors, especially at a genome-wide level. Identifying genetic factors related to BI may help researchers better understand the mechanisms underlying poor responses to vaccination, shedding light on the pathogenesis of COVID-19. Also, the identified genetic factors may be useful for guiding drug repurposing in the future^9^.

Here, we conducted a genome-wide association study (GWAS) for breakthrough infection (BI) (COVID-19 BI) based on the UK Biobank (UKBB). To the best of our knowledge, there are no published works on GWAS of COVID-19 BI yet. This is likely the first GWAS to investigate the genetic basis of COVID-19 BI and severe infection (focusing on pre-Omicron variants), including a comparison of severe vs mild BI, coupled with detailed post-GWAS bioinformatics analyses. The workflow in our study was shown in Figure 1. Briefly, we defined different study cohorts according to the number of vaccine doses received and whether the participants developed hospitalized or fatal BI. Then we performed GWAS analysis based on each scenario to identify the underlying genetic loci. Post-GWAS analysis was also conducted, including gene-based, pathway enrichment, and transcriptome-wide association studies (TWAS) analyses, as well as polygenic risk score (PRS) association analysis with other related medical conditions.

**Figure 1.**
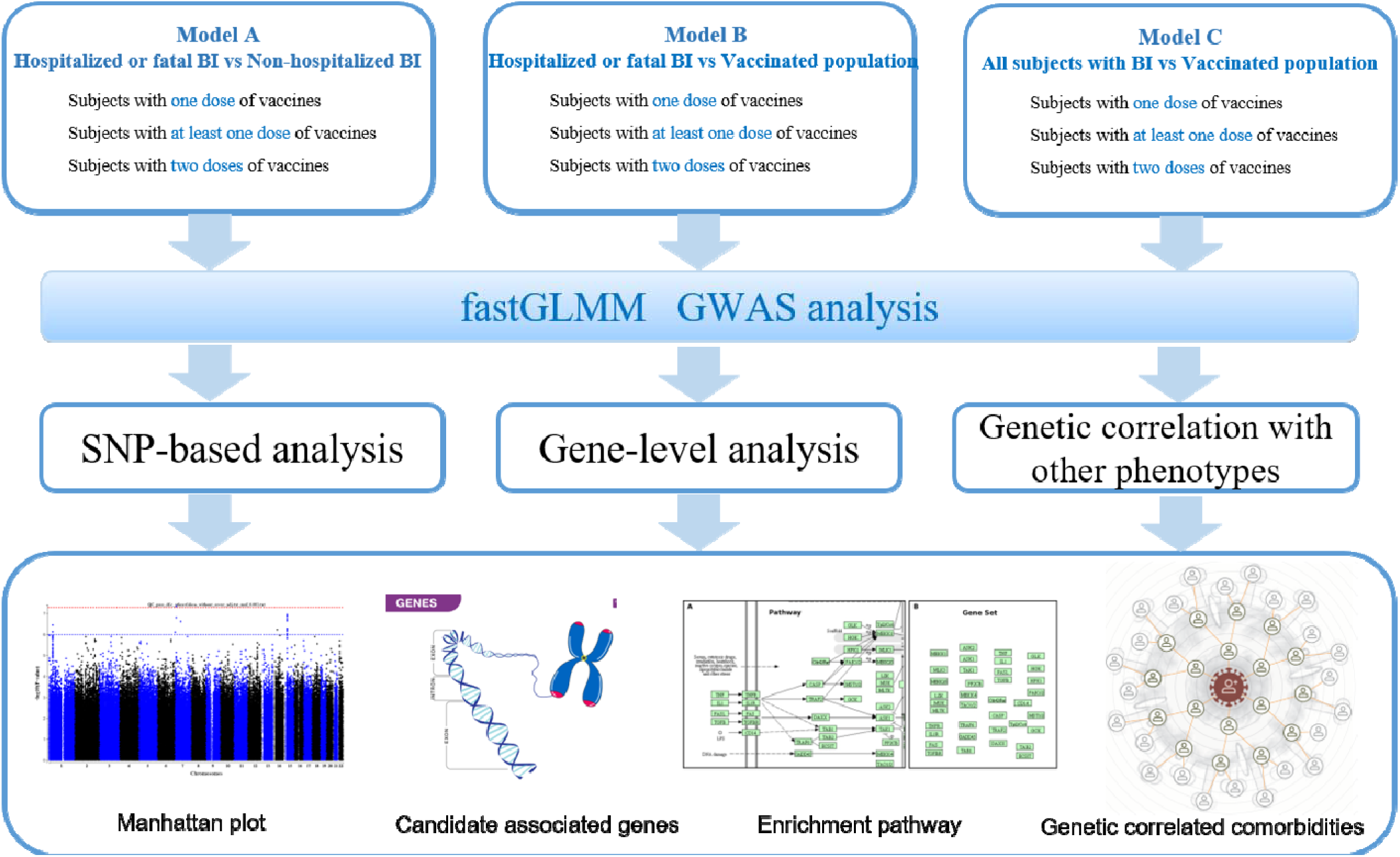
Workflow of our study

## Methods

### Participants and Cohort Definition

#### Data source

All the individual-level data in our study were extracted from the UK Biobank (UKBB), a large-scale prospective cohort comprising ∼500,000 individuals. The age of individuals in the current study varied from 50 to 89. Our current analysis was based on UKBB project number 28732^10^.

#### COVID-19 infection status

COVID-19 infection data were downloaded from the UKBB data portal. (for details, please refer to https://biobank.ndph.ox.ac.uk/showcase/exinfo.cgi?src=COVD19). Briefly, the latest COVID-19 test results were downloaded from UKBB, with the last update on 21 Jul 2021. COVID-19 infection status was primarily defined based on test results. Besides, COVID-19 diagnosis was also made based on ICD code U071 from hospital inpatient or mortality records, or code “Y2a3b” in TPP General Practice clinical records.

#### Vaccination status

Vaccination status was extracted from the TPP and EMIS GP clinical records (TPP last update 21 Jul 2021; EMIS last update 10 Aug 2021). Because the type of vaccine was missing in our datasets for most of the individuals, we did not perform analysis by vaccine type. Known data indicated participants received either BioNTech BNT162b2 or Oxford-AstraZeneca ChAdOx1 nCoV-19 vaccines (the median length of follow-up for the vaccinated group was 54 days). We defined three groups based on vaccination status: one dose, at least one dose, and two doses.

#### Inclusion and Exclusion criteria

Firstly, we included individuals with vaccination records under the TPP and EMIS systems (sample size *N*=393,544). Individuals with a prior infection were excluded as previous infections may also confer immunity. Afterwards, individuals with available imputed genotype data and labeled as European ancestry (UKB data-field 22006) were included.

#### Phenotype definition

COVID-19 BI was defined as an infection occurring 14 days after vaccination. If a subject received one dose of vaccination before the date of infection, we define this scenario as ‘one dose of vaccine’. The same applies to other dosages of vaccination.

We defined three cohorts A, B and C based on different criteria (Table 1). Cohort A compared hospitalized or fatal BI to non-hospitalized BI. Cohort B compared hospitalized or fatal BI to individuals without COVID-19 BI. Cohort C compared all BI cases to individuals without BI.

**Table 1.**
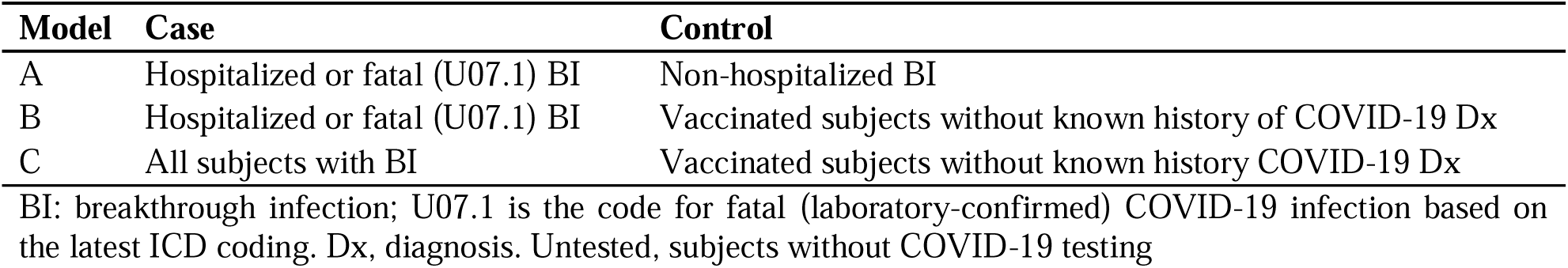
Definitions of models for covid-19 breakthrough infections.

### Genotyping and Quality Controls

Genotyping and data imputation were performed by the UKBB using Applied Biosystems UK BiLEVE Axiom Array (~□50,000 participants) and Applied Biosystems UK Biobank Axiom Array (~□450,000 participants)^11^. Marker positions were aligned to the GRCh37 reference genome.

In the first step, quality control (QC) of imputed genotyping data was performed by PLINK 1.9 to include a relatively small set of SNPs for computing the genetic relationship matrix (GRM). Briefly, we excluded SNPs with minor allele frequency (MAF) below 1%, minor allele count (MAC) below 100, genotype missingness above 10%, and Hardy-Weinberg equilibrium p-value less than 1e-10, and samples with more than 10% missingness. In total, 485,623 common variants with MAF > 0.01 and 488,371 individuals remained after the QC. These variants were used to compute the sparse genetic relationship matrix (GRM). Imputation was carried out by the UKBB (resulting in ~96M genotypes)^12,13^. Details are provided elsewhere (https://biobank.ndph.ox.ac.uk/showcase/showcase/docs/impute_ukb_v1.pdf).

The imputed data were filtered with standard QC criteria, e.g., MAC ≥ 10^14^, HWE test P ≥ 1e-10, genotyping rate ≥ 0.9, and imputation info score ≥ 0.3. The resulting set of imputed variants (ranging from 5,638,489 to 12,275,176 across cohorts) was used in the final GWAS analyses (Table S21).

### Genome-wide association study

GWAS was performed using a generalized linear mixed model (GLMM)-based method to test for association between imputed SNP dosages and BI phenotypes in cohort A, B and C. We employed fastGWA-GLMM^15^ to perform the GWAS analysis. This tool calculated a sparse genomic relationship matrix to evaluate pedigree-relatedness among individuals. In addition, fastGWA-GLMM can handle imbalanced data (for example when cases are rare compared to controls). We fitted age, sex, age*age, age*sex, and the top 10 genetic principal components provided by UKBB (data-field 22009) as covariates.

### SNP-based Analysis

LD-clumping was further performed using PLINK 1.9 (r^2^=0.5, distance = 250kb) to identify the independent loci. The European samples in Phase 3 1000 Genomes were used as the LD reference (GRCh37)^16^. SNP-to-Gene mapping was performed by the Bioconductor^17^ package ‘biomaRt’^18^ (version 2.48.2) on R-4.0.3. In addition, the OpenTargets Genetics portal^19^ was employed to prioritize the most relevant genes for each variant as a supplementary analysis.

### Gene-set and pathway analyses

#### Gene-based test with fastBAT

Gene-based test was performed using fastBAT^20^, with 1000 Genomes European ancestry samples as the LD reference^21^.

#### Multiple testing controlled by FDR

False discovery rate (FDR) was used to control for multiple testing. The Benjamini–Hochberg procedure (BH) adjusted P-value were used^22^. We set a FDR threshold of 0.05 to declare significance, while FDR<0.1 is considered an ‘suggestive’ association.

#### Pathway and Gene Ontology (GO) enrichment analyses with GAUSS^23^

Enrichment analysis of biological pathways was performed by Gene set analysis Association Using Sparse Signals (GAUSS)^23^.

Two collections of gene-sets (C2 and C5) were used, obtained from the Molecular Signature Database (MsigDB v6.2)^24^. C2 refers to a collection of curated pathways, including many canonical pathways such as KEGG, BioCarta, etc. C5 is another collection containing gene-ontology (GO) gene-sets. GAUSS identifies a subset of genes (called the core subset) within the gene set, which produces the maximum signal of association.

The corresponding p-value and core subset (CS) of genes for each outcome-pathway combination were computed via a composition of copula-based simulation and generalized pareto distribution (GPD)^25^. BH procedure for FDR control was used to correct for multiple testing.

### Transcriptome-wide association studies (TWAS) and Meta-TWAS

TWAS provides a novel approach for gene-trait association studies. TWAS utilizes known genetic variants (eQTLs) associated with transcript abundance to infer gene expression from GWAS data, thereby exploring associations between genetically regulated gene expression and complex traits. Here we performed TWAS for 48 tissues (see Table S17.1), including whole blood and lung tissues in GTEx v8 using the program S-PrediXcan^26^ FDR was used to correct multiple testing. We also performed a ‘meta-TWAS’ using S-Multixcan, integrates the results across different tissues to enhance statistical power^27^.

### Phenome Wide Association Studies

Phenome-wide association study (PheWAS) was performed to study the associations between SNPs and a large number of different phenotypes. We performed PheWAS via the OpenTargets Genetics portal^19^ with summary statistics from the UK Biobank, FinnGen, and GWAS Catalog.

### Evaluating genetic overlap of COVID-19 breakthrough infections with other medical conditions

#### Polygenic risk score analysis

In order to explore genetic overlap of COVID-19 BI with other conditions, we performed polygenic risk scores (PRS) analyses based on summary statistics using ‘PRsice’^28^. The summary statistics GWAS data were obtained from FinnGen^29^ and included a variety of medical conditions such as asthma, heart failure, cardiovascular diseases, obesity, diabetes, etc. (Table S18). Here we employed FinnGen maily to ensure no overlap with our UKBB samples. Different p-value thresholds (from 5e-8 to 0.01) were explored to filter the SNPs in PRS analysis. LD-clumping was performed at *r*^2^=0.05 within a distance of 250kb by PLINK 1.9. Harmonization of different sets of summary statistics was performed with ‘TwoSampleMR’ (version 0.4.26)^30^.

#### Genetic dependence between BI and other disorders using full GWAS summary statistics

Inspired by a recent study^31^, we also employed the Hoeffding’s test^32^ to evaluate genetic dependence across COVID-19 BI and other diseases. As demonstrated in the aforementioned study^31^, Hoeffding’s test of independence presents a viable alternative to LD score regression, particularly when dealing with small or moderate (effective) sample sizes, while maintaining adequate control of type I errors. (In this study, since the number of cases is in general limited, the effective sample size might be too small for a reliable LD score regression analysis.) In brief, Hoeffding’s test is a well-established non-parametric method that examines the marginal and joint distributions of two input variables (say *X* and *Y*) ^33^ and determines whether the distributions are independent. This test relies on the ranks of *X* and *Y*, avoiding parametric assumptions.

Our testing procedure closely mirrored that described in the reference^31^ and our recent study^34^. We performed clumping using PLINK (v1.9), setting the physical distance threshold at 10,000 kb and the *r*^2^ threshold at 0.2. We tested genetic dependence of COVID-19 BI with a range of other medical conditions, such as disorders of the respiratory, cardiovascular, endocrine and neurological systems (please refer to Table S18 for a comprehensive list). We utilized the R package ‘independence’ ^32^ to conduct the analysis.

## Results

### Results from SNP-Based Analysis

#### Results from GWAS

We performed GWAS analysis on 9 scenarios (Table 2). We identified two loci that were significantly associated with COVID-19 BI at the genome-wide level (p<5e-8), for ‘at least one dose of vaccine’ and ‘two doses of vaccine’ of cohort C (i.e., models C2 and C3, Table 3-4). The loci were rs36170929 on chromosome 7 (effect allele = G, effect size = 0.21, SE=0.038, allele frequency of G = 0.64, P=4.39e-8), and rs28645263 on chromosome 5 (effect allele = C, effect size = 0.35, SE=0.06, allele frequency of G = 0.42, P=9.46e-9).

**Table 2.**
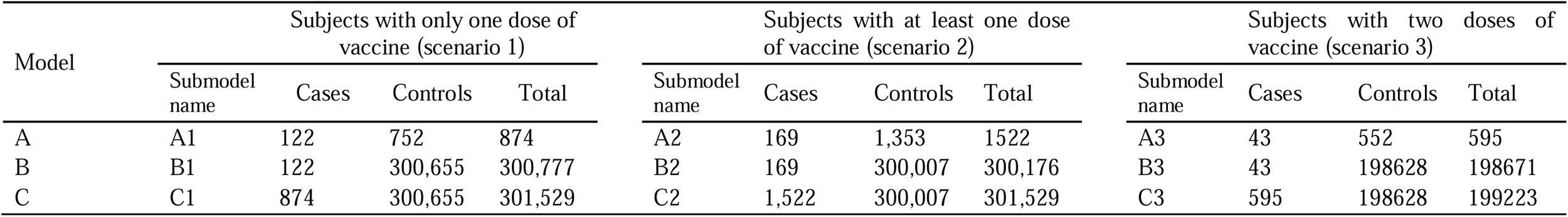
Number of available subjects of different models.

**Table 3.** Top10 SNP-based results of model C for participants with at least one dose of vaccine.

| SNP | Chr. | Location (bp) | Effect allele | Non-effect allele | Frequency of effect allele | BETA | SE | P | N | INFO | GeneSymbol | Gene name | Total no.of clumped SNPs | S0001 | Top gene prioritized by OpenTargets |
| --- | --- | --- | --- | --- | --- | --- | --- | --- | --- | --- | --- | --- | --- | --- | --- |
| rs36170929 | 7 | 12541187 | G | A | 0.640 | 0.210 | 0.038 | **4.39E-08** | 301529 | 0.984254 |  |  | 11 | 5 | VWDE |
| rs56150535 | 15 | 31647722 | T | C | 0.359 | 0.203 | 0.038 | 1.09E-07 | 301529 | 0.996787 | KLF13 | Kruppel like factor 13 | 33 | 20 | KLF13 |
| rs181987785 | 1 | 34977912 | G | A | 0.005 | 1.449 | 0.284 | 3.48E-07 | 301529 | 0.984316 |  |  | 30 | 30 | GJB5 |
| rs187268954 | 3 | 116529463 | C | T | 0.004 | 1.736 | 0.358 | 1.22E-06 | 301529 | 0.90264 |  |  | 4 | 3 | LSAMP |
| rs7590599 | 2 | 108915136 | C | T | 0.604 | 0.182 | 0.038 | 1.26E-06 | 301529 | 0.989482 | SULT1C2 | sulfotransferase family 1C member 2 | 8 | 5 | SULT1C2 |
| rs3737328 | 13 | 110866065 | T | C | 0.246 | 0.198 | 0.042 | 3.05E-06 | 301529 | 1 | COL4A1 | collagen type IV alpha 1 | 4 | 4 | COL4A1 |

| chain |  |  |  |  |  |  |  |  |  |  |  |  |  |  |  |
| --- | --- | --- | --- | --- | --- | --- | --- | --- | --- | --- | --- | --- | --- | --- | --- |
| rs142193221 | 22 | 21166165 | A | G | 0.006 | 1.274 | 0.274 | 3.31E-06 | 301529 | 0.929992 | *PI4KA* | phosphatidylinositol 4-kinase alpha | 6 | 6 | *PI4KA* |
| rs56070971 | 1 | 35025879 | T | C | 0.006 | 1.275 | 0.276 | 3.86E-06 | 301529 | 0.968667 |  |  | 29 | 29 | *GJB5* |
| rs72664942 | 4 | 85808904 | G | A | 0.007 | 1.174 | 0.259 | 5.75E-06 | 301529 | 0.938787 | *WDFY3* | WD repeat and FYVE domain containing 3 | 2 | 2 | *WDFY3* |
| rs79158353 | 10 | 78798475 | A | T | 0.082 | -0.304 | 0.067 | 6.48E-06 | 301529 | 0.995184 | *KCNMA1* | potassium calcium-activated channel subfamily M alpha 1 | 23 | 13 | *KCNMA1* |

1) S0001, number of clumped SNPs (SNPs in LD) with $p < 1e-3$ ; only SNPs with S0001 $\geq 2$ are shown.
2) LD clumping settings: $r^2 = 0.5$ , distance = 250kb

**Table 4.**
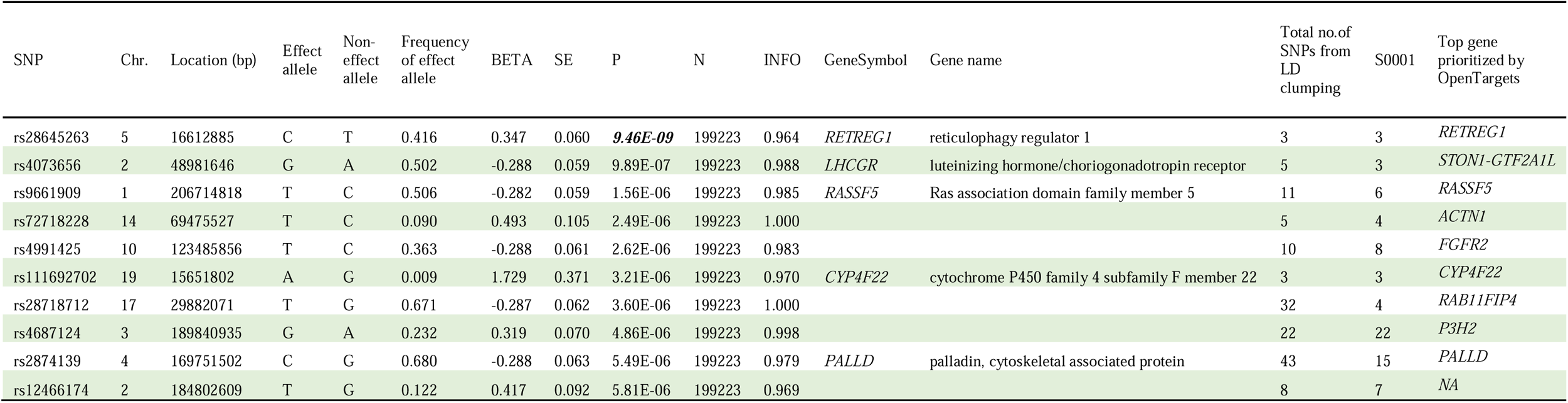
Top10 SNP-based results of model C for participants with two doses of vaccine.

Manhattan plots for GWAS of ‘at least one dose of vaccine’ and ‘two doses of vaccine’ are shown in Figures S1-2. Tables 3 and 4 show the top 10 SNPs found in models C2 and C3 for cohort C, respectively. All SNPs with p<1e-5 in the 9 scenarios are listed in Tables S1-9.

#### Significant SNPs mapped to genes

The rs36170929 locus maps to *LOC102725191*, an uncharacterized protein-coding gene. Based on the OpenTargets Genetics database, the top gene mapped to this SNP is *VWDE* (Von Willebrand Factor D And EGF Domains; distance to this gene = 97.62 kb), as rs36170929 is an eQTL for *VWDE*. The rs28645263 locus maps to *RETREG1* (Reticulophagy Regulator 1).

For the top 10 independent SNPs associated with COVID-19 BI in Tables 3-4, the most probable disease-associated genes corresponding to these SNPs were further prioritized by the OverallV2G (Variant-to-Gene) score from OpenTargets Genetics (Table S10). Additional assigned genes using OpenTargets Genetics for SNPs with GWAS p-value < 1e-4 are listed in Table S11.

#### Region plots of significant SNPs

Region plots of rs36170929 and rs28645263 were shown in Figure S3 and Figure S4, displaying LD-clumped SNPs with these significant loci located within 1Mb.

### Results from Gene-Based Analysis

#### Results of fastBAT

We employed fastBAT to perform further gene-level analysis, focusing on common variants (MAF>0.01). Top 10 genes from the gene-based analyses are listed in Table 6. The gene *BAGE* (P=3.86e-8, FDR = 9.51e-4, chromosome 21) reached significance (FDR < 0.05) after adjusting the p-value by the BH procedure, while genes *BAGE2, BAGE3, BAGE4, BAGE5, ARHGEF3* were considered having suggestive associations with BI with FDR < 0.1 (Table S12).

#### Results of pathway enrichment analysis by GAUSS

To gain deeper insights into the relevant functional pathways, we employed GAUSS for further analysis of genes extracted from fastBAT. Totally 10,679 canonical pathways and gene ontology (GO) gene sets from the MSigDB database were tested.

Table 5 shows the pathway enrichment analysis results. For the results of canonical pathways, some of the top enriched pathways included KEGG VIRAL MYOCARDITIS (FDR corrected p = 0.05), BIOCARTA AKAP13 PATHWAY (FDR corrected p = 0.06), KEGG TIGHT JUNCTION (FDR corrected p = 0.06), and REACTOME TRANSLATION (FDR corrected p = 0.06). More detailed results are listed in Table S13-14. For the results of GO gene sets (C5), the top significant associations were observed based on Model A (participants with at least 1 dose of vaccine) for GOCC MUSCLE MYOSIN COMPLEX (FDR corrected *p* = 1.44e-5), GOCC MYOSIN FILAMENT (FDR corrected *p* = 1.44e-5), and GOCC MYOSIN COMPLEX (FDR corrected *p* = 6.41e-4).

**Table 5.**
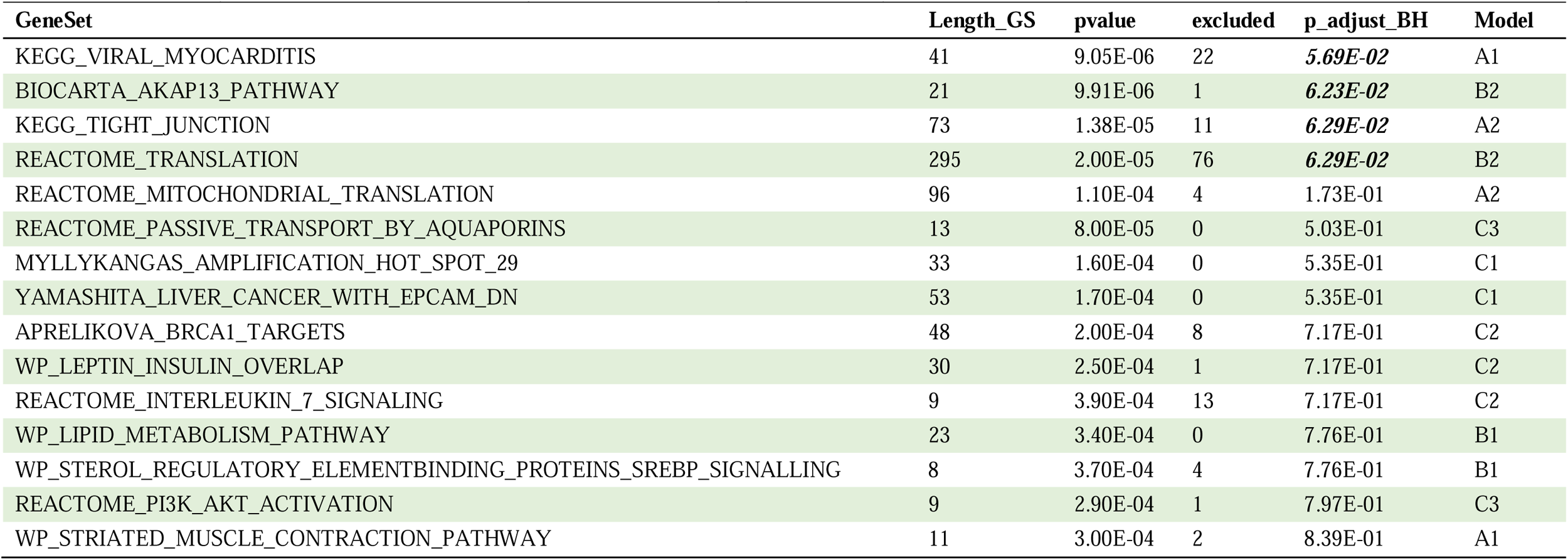
Top 15 pathway enrichment results (GAUSS) for genes identified through gene-based analysis (fastBAT)

**Table 6.**
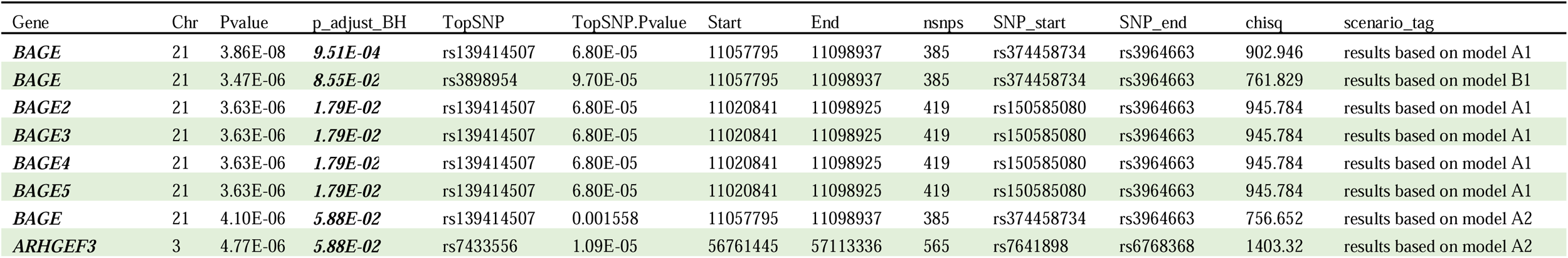
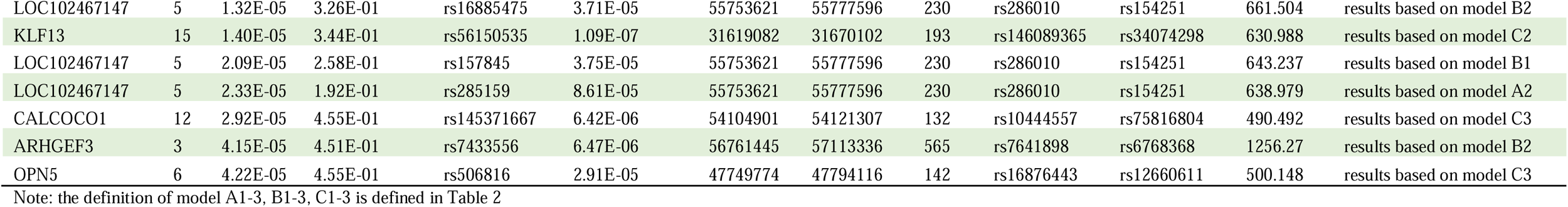
Top15 results of gene-based analysis based on all the model in our study.

#### Results from TWAS

We employed S-Multixcan to investigate the associations between genetically regulated gene expression and phenotypes across 48 types of human tissues (TableS17.1), and combine evidence across these tissues to improve statistical power. The most significant association with COVID-19 BI was observed for *AQP7P1* (FDR corrected P = 7.34e-3). Further, *PFN1P2* (FDR corrected P = 1.61e-2), *AL590452.1* and *LINC00842* (FDR corrected P <0.05) were observed to be associated. In addition, *RP11-314D7.3* (FDR corrected P=6.94e-2) showed moderate associations with BI (FDR between 0.1 and 0.2). More results are provided in Table S17.2.

### Results from analysis of genetic overlap with other conditions

#### Results of PRS and genetic dependence analysis of breakthrough infection with other medical conditions

We performed polygenic risk score testing for BI with other medical conditions to explore polygenic associations. Table 7 lists the results based on model C2 for individuals with at least one dose of vaccine. The most significant positive association was observed for heart failure (FDR corrected P = 1.82e-3). We also observed significant associations of BI with HbA1c (FDR corrected P = 2.18e-2), and type I diabetes (FDR corrected P = 1.22E-02). We also found nominally significant associations (nominal p-value <0.05) for several traits such as obesity, BMI, dementia, asthma, COPD/asthma-related infections, serum urate etc. (Table S19).

**Table 7.** Polygenic association testing of BI (model C2, general BI vs population) with related traits using summary statistics (p<0.05 are shown)

| Body system | Exposure | pval_PRS | p_adjust_BH | coefficient | r2 | nsnps | exposure_p_filter | clump_r2 |
| --- | --- | --- | --- | --- | --- | --- | --- | --- |
| cardiovascular system | Heart Failure | **1.33E-04** | **1.82E-03** | 0.030588 | 4.84E-05 | 41900 | 0.05 | 0.05 |
| endocrine system | Type 1 diabetes, strict (exclude type 2) | **1.00E-03** | **1.22E-02** | 0.028586 | 3.59E-05 | 131 | 5.00E-08 | 0.05 |
| endocrine system | Glycaemic_HbA1c | **1.96E-03** | **2.18E-02** | 0.704835 | 3.18E-05 | 250 | 1.00E-04 | 0.05 |
| endocrine system | Diabetes mellitus (type 1 and 2) | **1.61E-02** | 1.30E-01 | 0.097242 | 1.92E-05 | 128 | 5.00E-08 | 0.05 |
| endocrine system | Obesity | **3.63E-02** | 2.37E-01 | 0.004535 | 1.45E-05 | 56424 | 0.05 | 0.05 |
| endocrine system | BMI | **1.49E-02** | 1.32E-01 | 0.17485 | 1.97E-05 | 1365 | 1.00E-07 | 0.05 |
| immune system | Human immunodeficiency virus disease | **3.71E-02** | 2.41E-01 | 0.042006 | 1.44E-05 | 17 | 1.00E-05 | 0.05 |
| nervous system | Dementia | **2.95E-02** | 2.00E-01 | 0.003864 | 1.57E-05 | 78932 | 0.1 | 0.05 |
| respiratory system | COPD/asthma related infections | **9.15E-03** | **8.58E-02** | 0.01321 | 2.25E-05 | 54680 | 0.05 | 0.05 |
| respiratory system | Asthma | **2.09E-02** | 1.62E-01 | -0.009499 | 1.77E-05 | 20426 | 0.01 | 0.05 |
| respiratory system | Smoking Cessation | **4.00E-02** | 2.46E-01 | 0.15793 | 1.40E-05 | 2871 | 0.001 | 0.05 |
| renal system | Diabetic kidney disease in type 1 DM | **9.75E-03** | **9.00E-02** | -0.015166 | 2.22E-05 | 1449 | 0.001 | 0.05 |
| renal system | Serum urate | **1.16E-02** | 1.04E-01 | 1.205126 | 2.11E-05 | 33 | 0.05 | 0.05 |

\*(1) clump\_r2=0.05. (2) More details about the information for each exposure are listed in Table S18
(2) All the outcome in this table is Model C2 defined in Table 2

Also, we performed Hoeffding’s independence test to evaluate genetic dependence between these comorbidities and BI. Table 8 and Table S20 show the results of Hoeffding’s Independence test of BI with related traits for individuals with at least one dose of vaccine. Several conditions including asthma, abnormal findings on lung imaging, type I diabetes and schizophrenia showed significant genetic dependence with FDR<0.05, while a few other traits including pulmonary embolism and cardiomyopathy showed FDR<0.1. A variety of other pulmonary, cardiometabolic, neurological and liver conditions were nominally significant at p<0.05.

**Table 8.** Hoeffding’s Independence test of BI with related traits using summary statistics (p<0.05 are shown)

| Exposure | Outcome | pthres | n | Dn | scaled | p.value | p.adj_pthres&traitB_sepa<br>rate |
| --- | --- | --- | --- | --- | --- | --- | --- |
| **Respiratory** |  |  |  |  |  |  |  |
| Abnormal findings on diagnostic imaging of lung | A2 | 0.1 | 102776 | 1.29E-06 | 4.76 | **2.05E-04** | **8.00E-03** |
| Abnormal findings on diagnostic imaging of lung | B2 | 0.1 | 102787 | 7.19E-07 | 2.66 | **4.47E-03** | 1.74E-01 |
| Asthma (only as main-diagnosis) | A2 | 0.5 | 372099 | 1.94E-07 | 2.6 | **4.88E-03** | 1.43E-01 |
| Asthma (only as main-diagnosis) | B2 | 0.5 | 372141 | 1.81E-07 | 2.42 | **6.41E-03** | 8.33E-02 |
| Asthma (only as main-diagnosis) | C2 | 0.05 | 68429 | 1.55E-06 | 3.82 | **8.08E-04** | **2.69E-02** |
| Asthma, hospital admissions, main diagnosis only | A2 | 0.5 | 371828 | 1.63E-07 | 2.19 | **9.11E-03** | 1.43E-01 |
| COPD/asthma related infections | B2 | 1.00E-05 | 44 | 8.24E-04 | 1.28 | **3.77E-02** | 2.56E-01 |
| COPD/asthma related pneumonia or pneumonia derived septicaemia | A2 | 0.01 | 15042 | 2.35E-06 | 1.27 | **3.81E-02** | 2.97E-01 |
| Interstitial lung disease | A2 | 0.3 | 248253 | 2.02E-07 | 1.81 | **1.63E-02** | 3.08E-01 |
| Interstitial lung disease endpoints | C2 | 0.2 | 190993 | 1.70E-07 | 1.17 | **4.49E-02** | 6.65E-01 |
| Obesity related asthma | A2 | 0.01 | 15347 | 3.12E-06 | 1.73 | **1.85E-02** | 2.41E-01 |
| Obesity related asthma | B2 | 0.01 | 15350 | 2.12E-06 | 1.17 | **4.48E-02** | 5.82E-01 |
| Pulmonary embolism | B2 | 0.05 | 58577 | 1.46E-06 | 3.07 | **2.43E-03** | 6.17E-02 |
| Tuberculosis | A2 | 0.01 | 13123 | 3.08E-06 | 1.45 | **2.84E-02** | 2.77E-01 |
| **Cardiovascular** |  |  |  |  |  |  |  |
| Cardiomyopathy | C2 | 0.1 | 103175 | 9.18E-07 | 3.41 | **1.48E-03** | 5.75E-02 |
| Cardiomyopathy (excluding other) | B2 | 0.5 | 363183 | 1.87E-07 | 2.44 | **6.18E-03** | 8.33E-02 |
| Cardiomyopathy (no controls excluded) | A2 | 0.01 | 14204 | 4.44E-06 | 2.27 | **8.07E-03** | 1.57E-01 |
| **Endocrine** |  |  |  |  |  |  |  |
| Diabetes mellitus (type 1 and 2) | A2 | 0.3 | 275918 | 1.17E-07 | 1.16 | **4.52E-02** | 3.52E-01 |
| Diabetes mellitus (type 1 and 2) | C2 | 0.1 | 129409 | 4.66E-07 | 2.17 | **9.33E-03** | 1.82E-01 |
| Obesity | B2 | 0.4 | 319934 | 1.34E-07 | 1.54 | **2.49E-02** | 3.95E-01 |
| Type 1 diabetes, strict definition | A2 | 1.00E-04 | 728 | 9.35E-05 | 2.45 | **6.16E-03** | 1.20E-01 |
| Type 1 diabetes, wide definition | B2 | 0.2 | 179971 | 6.59E-07 | 4.27 | **4.21E-04** | **1.64E-02** |
| Type 1 diabetes, wide definition | C2 | 0.05 | 56637 | 6.89E-07 | 1.41 | **3.07E-02** | 3.99E-01 |
| **Neurological** |  |  |  |  |  |  |  |
| Schizophrenia or delusion | C2 | 1.00E-05 | 35 | 2.00E-03 | 2.44 | **6.19E-03** | 2.41E-01 |
| Schizophrenia or delusion (more controls excluded) | A2 | 0.01 | 15032 | 6.43E-06 | 3.48 | **1.33E-03** | 5.20E-02 |
| Schizophrenia, schizotypal and delusional disorders | B2 | 1.00E-05 | 43 | 3.09E-03 | 4.68 | **2.33E-04** | **9.09E-03** |
| Any dementia | B2 | 1.00E-05 | 109 | 4.62E-04 | 1.8 | **1.66E-02** | 2.56E-01 |
| Any dementia (more controls excluded) | A2 | 0.001 | 1858 | 2.18E-05 | 1.45 | **2.84E-02** | 2.77E-01 |
| **Liver** |  |  |  |  |  |  |  |
| Alcoholic liver disease | A2 | 0.001 | 1690 | 3.69E-05 | 2.25 | **8.35E-03** | 2.77E-01 |
| Cirrhosis, broad definition | A2 | 1.00E-04 | 202 | 3.92E-04 | 2.84 | **3.43E-03** | 1.20E-01 |
| Cirrhosis, broad definition | C2 | 0.3 | 248811 | 1.36E-07 | 1.22 | **4.12E-02** | 6.57E-01 |
| Nonalcoholic fatty liver disease | B2 | 0.2 | 178801 | 1.82E-07 | 1.17 | **4.45E-02** | 4.34E-01 |

1) More details about the information for each exposure are listed in Table S18.
2) Scaled statistic: the test statistic rescaled for a standard null distribution (please refer to the R package “independence” for details). FDR adjusted-p < 0.05 are in bold and those between 0.05 and 0.1 are in italics. FDR adjustment was performed with stratification by trait B
3) r2 threshold for LD-clumping is 0.2
4) The definition of the outcomes is listed in Table 2

#### Results of PheWAS with the top associated variants

The PheWAS results for the top 10 SNPs identified in Models C2 and C3, based on individuals receiving at least one or two doses of the vaccine, revealed several SNPs significantly associated with lymphocyte counts and white blood cell counts. Although some did not reach genome-wide significance (P = 5e-8).

Specifically, rs28645263 (P = 3.60e-4, Beta = 0.0078) and rs9661909 (P = 2.64e-6, Beta = −0.008922) were significantly associated with lymphocyte counts in PheWAS, with corresponding GWAS P-values of 9.46e-9 and 1.56e-6, respectively. Additionally, rs28645263 (P = 9e-4, Beta = 0.0073) and rs4073656 (P = 1.23e-5, Beta = 0.008) were associated with white blood cell counts, with GWAS P-values of 9.46e-9 and 9.89e-7, respectively. Further details are provided in Tables S15-16.

## Discussion

In this study, we conducted a GWAS study to uncover the associated genetic factors of BI using data from the UKBB. Furthermore, a series of post-GWAS analysis, including gene-based analysis, pathway enrichment analysis, PRS analysis etc., were performed to unveil new insights into the genetic architecture of BI. To the best of our knowledge, this is the first GWAS to investigate the genetic basis of breakthrough COVID-19 infection (BI) and severe infection (focusing on pre-Omicron variants), including a comparison of severe vs mild BI.

### Interpretation of findings

#### Top loci identified from GWAS

We identified two loci, rs36170929 (*p*=4.39e-8) and rs28645263 (*p*=9.46e-9), which showed association with COVID-BI at genome-wide significance. These two loci can be mapped to two protein-coding genes, *LOC102725191* and *RETREG1* (Reticulophagy Regulator 1) respectively. *RETREG1* is widely considered as an important mediator of reticulophagy (also referred as ER-phagy). Reticulophagy is a specific type of autophagy which involves the selective elimination of portions of the endoplasmic reticulum (ER)^35^. Notably, a recent study^36^ found that the ER-associated degradation (ERAD) regulator ERLIN1 strongly impeded the late stages of SARS-CoV-2 replication. Furthermore, it was discovered that two additional factors, *RETREG1* and *FNDC4*, which are involved in ER-phagy and aggresome-related processes respectively, also hindered SARS-CoV-2 replication. These findings suggest that components of the ERAD pathway, including RETREG*1,* may serve as inhibitors of COVID-19 infection. However, the precise mechanisms by which this gene influences COVID-19 BI warrant further investigation.

Although *LOC102725191* is a protein-coding gene, its function remains uncharacterized. Based on OpenTargets, another gene *VWDE* (Von Willebrand Factor D And EGF Domains) was listed as the top gene mapped to rs3617092, as this SNP is an eQTL for *VWDE*. Von Willebrand Factor (vWF) is a multimeric glycoprotein that is involved in inflammation and hemostasis. It has been reported that COVID-19 is associated with elevated levels of vWF antigen and activity, which may be linked to an increased risk of thrombosis in infected patients^37^.

As for the other top loci, a study^38^ showed that Kruppel-like factor 13 (*KLF13*) has low activity in moderate COVID-19 patients and high activity in severe cases. Low *KLF13* expression is associate with reduced pro-inflammatory and enhanced phagocytic activity in macrophages, necessary for an efficient immune response^39^. These results support *KLF13*’s association with COVID-19 severity^40^.

#### Gene-based results

Several *BAGE* family member genes, including *BAGE, BAGE2, BAGE3, BAGE4, BAGE5*, were observed to be significantly associated with BI in the gene-based analysis. *BAGE* (B Melanoma Antigen) is a protein-coding gene. This gene encodes a tumor antigen recognized by autologous cytolytic lymphocytes (CTL)^41^. There is currently no direct literature or study to support the association between BAGE and COVID-19 or related diseases yet, and further studies are needed. In addition, we also observed *ARHGEF3* was suggestively associated with BI. In another bioinformatics analysis^42^ of differentially expressed genes targets in SARS-CoV-2, *ARHGEF3* reached significance (P.adjust = 0.002415, table 1 of ref^42^), yet further validation studies are required.

#### Pathway and GO enrichment analysis

The most significant result in our pathway enrichment analysis was related to KEGG VIRAL MYOCARDITIS. Viral myocarditis is a cardiac disease associated with inflammation and injury of the myocardium. Myocarditis may be caused by direct cytopathic effects of the virus, a pathologic immune response to persistent virus, or autoimmunity triggered by the viral infection. Of note, viral myocarditis is associated with both COVID-19 infection and vaccination. According to a study in Isreal, COVID-19 vaccination increased the 42-day risk of myocarditis by a factor of 3.24 (95% CI, 1.55 to 12.44) as compared to unvaccinated persons, with events mostly concentrated among young males^43^. On the other hand, COVID-19 itself is also linked to a significantly elevated risk of myocarditis^44^. It is intriguing that viral myocarditis is identified as the top-ranked pathway, which may suggest that the genes involved in myocarditis are also associated with immunological responses to vaccination. The core subset of genes identified by GAUSS in this pathway could be a focus for further experimental studies, potentially providing new insights into associations between COVID-19 BI and myocarditis.

Another pathway that also shows suggestive association with BI is the BIOCARTA AKAP13 PATHWAY (Rho-Selective Guanine Exchange Factor AKAP13 Mediates Stress Fiber Formation). The A-kinase anchor protein 13 (AKAP13, also known as AKAP-LBC) is a group of structurally diverse proteins, which have the common function of binding to the regulatory subunit of protein kinase A (PKA) and confining the holoenzyme to discrete locations within the cell^45^. A polymorphism near the *AKAP13* gene, associated with higher levels of *AKAP13* mRNA expression in the lung, has been reported to associate with higher risks of developing idiopathic pulmonary fibrosis (IPF)^46^. Several studies^47,48^ have shown positive and significant genetic correlation between IPF and COVID-19. In addition, AKAP13 has been shown to regulate Toll-like receptor 2 (TLR2) signaling and play a role in innate immune responses downstream of TLRs^49^.

It is also worth noting that lipid-related pathways are also ranked among the top, such as “WP_LIPID_METABOLISM_PATHWAY” and “WP_STEROL_REGULATORY_ELEMENTBINDING_PROTEINS_SREBP_SIGNALLING”. Sterol regulatory element-binding protein (SREBPs) are key regulators of lipid metabolism including synthesis of cholesterol^50^. During viral infection, lipids play a crucial role in various processes such as membrane fusion, replication, and endocytic and exocytic processes. Drugs targeting lipid metabolism has been suggested as drug targets as well^51,52,53^.

In line with our findings that PRS of diabetes-related traits are significantly associated with BI, the pathway leptin-insulin signaling overlap was also top-ranked. Obesity is a well-known risk factor for severe COVID-19 infection, although the mechanism remains unclear. It has been postulated that leptin, which regulates both appetite and immunity^54^, may contribute to the pathogenesis of COVID-19.

Interleukin-7 signaling pathway was also among the top pathways. Interleukin-7 (IL-7) is a cytokine crucial for T cell development and homeostasis. IL-7 has been studied as a potential therapeutic to treat severe COVID-19 patients with lymphopenia and lymphocyte exhaustion^55^.

Another enriched pathway was related to aquaporin signaling. Aquaporins are water channels that play a role in fluid homeostasis, and have been implicated in the development of pulmonary edema in respiratory diseases^56^. Another study showed that aquaporin levels were significantly elevated in critical COVID-19 patients^57^.

#### Polygenic score analysis and genetic overlap with other disorders

In the PRS association analysis, we observed a positive significant genetic association between COVID-19 BI with several traits, including heart failure and glycaemic traits (HbA1c) (FDR<0.05). A recent study also observed a positive genetic association between COVID-19 and heart failure^58^. Combined with our current findings, these results provided evidence to support shared genetic etiology between COVID-19 BI and heart failure. Heart failure has also been reported to be associated with more severe infections and as one of the long-term sequelae of COVID-19^59^.

In addition, our results showed a statistically significant association between HbA1c and COVID-19 BI. Interestingly, a related study^60^ showed that poor glycaemic control, assessed by mean HbA1c in the post-vaccination period, was associated with lower immune responses and an increased incidence of SARS-CoV2 BI in type 2 DM patients, consistent with our findings based on genetic data. Of note, we also observed significant genetic overlap of COVID-19 BI with type I diabetes, using both PRS analysis and genetic dependence analysis with Hoeffding’s test. A recent review summarized current studies on vaccine response and diabetes, with most studies reporting lower antibody response in diabetic patients^61^, and some studies reported that higher BMI may also be associated with poorer immunogenicity. However, the high heterogeneity and modest sample sizes of many studies preclude a firm conclusion from being made.

A range of cardiometabolic traits were also nominally significant in our PRS or genetic dependence analysis, although not passing the FDR correction. For example, obesity, BMI, diabetes mellitus (type I and II), and serum urate were observed to be have genetic overlap with BI. As discussed above, several pathways related to lipid metabolism, leptin-insulin signaling overlap etc. were among the top enriched ones. Taken together, our results may suggest that cardiometabolic traits share genetic bases with COVID-19 BI. As such, it will be intriguing to study whether these cardiometabolic traits are risk factors or complications of COVID-19 BI.

In the genetic dependence analysis with Hoeffding’s test, we observed several traits showing significant results passing FDR correction (FDR<0.05), including asthma, abnormal findings on diagnostic imaging of lung, schizophrenia, and type I diabetes. Given the possible genetic overlap between these traits and BI, these traits may be associated with increased risks of BI, or present as sequelae post-infection. However, further studies are necessary to elucidate these relationships.

### Strengths and limitations

Firstly, to the best of our knowledge, this is the first GWAS to investigate the genetic basis of breakthrough COVID-19 infection (BI) and severe infection (focusing on pre-Omicron variants), including a comparison of severe vs mild BI. Secondly, we conducted a comprehensive series of post-GWAS analysis to provide insights into the biological basis of COVID-19 BI. These include standard SNP-based tests as well as gene-based (fastBAT, S-MulTiXcan) and pathway-based (GAUSS) analyses, which may help bridge the gap between significant SNPs detected and the corresponding biological mechanisms. Lastly, we explored the genetic associations between COVID-19 BI and related disorders through PRS and other analyses.

Our study also has a few limitations. Firstly, although the total sample size in our study is large, the number of cases is relatively limited, due to a relatively short follow-up duration (maximum 253 days between vaccination and infection dates). However, studies^62^ have shown that vaccine effectiveness in preventing infection wanes over time^63^. This challenge makes it harder to capture specific genetic factors underlying vaccine response as follow-up length increases. We aimed to balance follow-up length and vaccine effectiveness to uncover the genetics of BI. Additionally, the UK Biobank population may not fully represent the entire UK population, as participants tend to be healthier and have higher socioeconomic status^64^ compared to non-participants. Furthermore, our study is based on European samples, and the generalizability of these genetic findings to other populations remains uncertain. Further studies in other populations are warranted.

In summary, we have conducted a GWAS for breakthrough infection with SARS-CoV-2 in a European population using UK Biobank data. A series of post-GWAS analysis was performed, including gene-based analysis, pathway enrichment analysis, PRS association, and others. We discovered two novel genetic loci and revealed corresponding genes and pathways that may underlie COVID-19 BI. We believe this work provides an important foundation and reference for future studies at elucidating the biological and genetic basis of COVID-19 breakthrough infections.

## Data Availability

All data produced in the present study are available upon reasonable request to the corresponding author, HCS

## Author contributions

Y.F designed and implemented the investigations, contributed to the analyses of the data and wrote the paper. CY. W provided suggestions on the methods, results, discussion, and revised the sections accordingly. WK. T performed part of the GWAS analysis. RY. Z helped with the Hoeffding’s D Independence Test. Y. X extracted the original data of BI from UKBB. HC. S conceived and supervised the study, contributed to methodology development and interpretation of results, and revised the paper. All authors reviewed, edited and approved the final paper.

## Acknowledgments

This work was supported partially by a National Natural Science Foundation China grant (81971706), a National Natural Science Foundation China (NSFC) Young Scientist Grant (31900495), the Lo Kwee Seong Biomedical Research Fund from The Chinese University of Hong Kong and the KIZ-CUHK Joint Laboratory of Bioresources and Molecular Research of Common Diseases, Kunming Institute of Zoology and The Chinese University of Hong Kong, China. We would like to thank Prof. Yang Jian and Dr. Jiang Longda for their great suggestions on technical problems of GCTA-GLMM. We would also like to thank Prof TSUI Kwok Wing Stephen and Prof. Cao Qin for useful discussions. We also thank Dr. Yin Liangying, Dr. SHI Yujia, Dr. Xue Xiao and Mr. Lin Yu-Ping for their advice on technical problems. An earlier version of this study was released as a preprint (http://dx.doi.org/10.13140/RG.2.2.25986.66248) on 30 Dec 2023.

## Supplementary Material

All supplementary files are available at https://drive.google.com/drive/folders/1ux1b3VK2NxnkVFVkowO68h5_AjhDz-xg?usp=drive_link

